# Olfactory detection of human odorant signatures in Covid patients by trained dogs

**DOI:** 10.1101/2021.07.12.21258827

**Authors:** Ing. Lenka Vlachová, Gustav Hotový, Ing. Jiří Šlechta, Ing. Roman Váňa, Bc. Milena Vokřálová, Mgr. Jiří Zeman

**Affiliations:** Search and Rescue Czech Republic

**Keywords:** COVID, detection, dog

## Abstract

The objective of the study was to verify the ability of specially trained dogs to detect the odour of people ill with COVID-19 and, at the same time, to use the outcome of this research in the future, whether in combatting a similar pandemic or in the field of medicine in the shape of a biological detector in uncovering different diseases. Our key assumption was that the disease will change the active odour signature of the individuals just like other diseases (TBC, malaria, tumours, etc.). The pilot study was conducted in two places, based on the same protocolar methods, and it included four specially trained detection dogs in total. For the first phase of the project, we obtained 156 positive and 72 negative odour samples primarily from a hospital. Each detection dog involved in the study was imprinted with the smell samples of Covid-positive people. The first experiment only involved two dogs. With the other two dogs, the phase of imprinting a specific smell was longer, possibly because these dogs were burdened with previous training. During a presentation of 100 randomised positive samples, the experimental dogs showed a 95% reliability rate. Data from this pilot study show that specially trained dogs are able to detect and identify the odour samples of people infected with the SARS-CoV-2 coronavirus.

## 1 Introduction

The new SARS-CoV-2 coronavirus appeared in Wuhan, China in December 2019. It triggered a pandemic of a serious human respiratory syndrome (COVID-19). The exponential global spread of the SARS-CoV-2 virus crimped the world with its massive socio-economic impact and its impact on public health. As of the end of 2020, more than 40 million people in 213 countries of the world had been infected and more than a million people had died worldwide. Although SARS-CoV-2 is the seventh known coronavirus type capable of infecting humans, the therapy is still not 100% efficient.

SARS-CoV-2 is a part of the family of β coronaviruses. Four of these coronaviruses (229E, NL63, OC43 and HKU1) cause only moderate symptoms of a cold. On the other hand, the other three, SARS-CoV, MERS-CoV and SARS-CoV-2, can cause serious symptoms and even death with mortality rates of 10 %, 37 % and 5 %, respectively (Zhu et al., 2020). SARS-CoV-2 is a single-stranded RNA virus. The metagenomic sequencing approach has been used to characterise the entire genome with a total length of 29,881 base pairs (GenBank no. MN908947) and encoding 9,860 aminoacids. Gene fragments express structural and non-structural proteins (Chen et al., 2020).

SARS-CoV-2 spreads primarily through respiratory globules during a face-to-face contact. The infection may spread by means of asymptomatic, presymptomatic and symptomatic carriers. The average time between exposure and the onset of symptoms is five days. The most frequent symptoms are fever, dry cough and breathing difficulties. SARS-CoV-2 is diagnosed through detection by means of a reverse transcription of polymerase chain reaction tests. False negative test results may occur with 20-67% of patients, depending on the quality, method and timing of the tests (Wiersinga et al., 2020).

To activate the entry factors, respiratory viruses such as influenza, parainfluenza and coronaviruses rely on host proteases, which facilitate the membrane fusion and an entry into the epitelial cells of the respiratory system. Transmembrane protease serine 2 is a ubiquitously expressed serine protease, which is crucial for the splicing and activation of both human influenza hemagglutinin and the spike (S) proteins of the SARS coronaviruses (Hoffmann et al., 2020). The angiotensin-coverting enzyme 2 (ACE2) is crucial for binding the SARS-CoV-2 virus to the surface of the host cell. The transmembrane serine protease activates the S protein and hence mediates the entry into the cell. When the virus enters the cell, virus RNA is released. The replication and transcription of the virus RNA genome take place through protein splicing and the assembly of a replicase – transcriptase complex. Virus RNA is replicated and structural proteins are synthesised, assembled and wrapped in the host cell into which virus particles are being released (Fehr et al.,2015).

The ongoing global pandemic of the SARS-CoV-2 coronavirus causing the COVID-19 disease has required an answer to the question whether it would be possible to use a specially trained dog as a biological detector. The objective of the study was to verify the ability of trained dogs to detect the COVID-19 disease in the population. Studies published earlier have shown dogs are able to detect various diseases.

Dogs have an extremely sensitive smell sense with a proved lower detection limit in concentrations of 10-12 of the total (ppt), which is three orders more sensitive than the currently available detection means, which can reliably detect substances in concentrations of up to one millionth (ppm) or one billionth of the total (ppb). To illustrate the extreme sensitivity of the canine sense of smell, a dog could detect the equivalent of a single drop in twenty Olympic swimming pools (Angle et al., 2016). The hypothesis that dogs might be able to detect malign tumours based on their specific smell was proposed in a clinical study as early as in 1989 (Williams, Pembroke). Detection dogs are also used as a bio-sensor as they can warn against an approaching epileptic or diabetic fit. It has been proved that dogs are able to detect bacteriological infections (Mauer, 2016). In Mauer’s study, dogs were trained to detect urine samples positive for the *E. Coli, Enterococcus, Staphylococcus aureus* and *Klebsiella* bacteria. These dogs were able to compare urine positive and negative for the bacteria. As regards the detection of viruses by specially trained dogs, it has been proved that dogs are able to distinguish cell cultures infected and uninfected with the bovine herpes virus or with bovine parainfluenza (Angle et al., 2016). Compared with bacteria, viruses do not have a metabolism of their own, and therefore the volatile organic compounds (VOC) are released by infected body cells as a result of metabolic host processes (Amann et al., 2014). VOC are compounds with a low molecular weight, which evaporate easily under normal temperatures and pressure. Volatile compounds can be found in exhaled breath, in the skin, urine, saliva, blood and excrements. VOC are released in concentrations ranging from ppb to ppt in human breath and ppm to ppb in human blood and urine (Schmidt et al., 2015). It has been proved that dogs detect amyl acetate in the ppt range (Walker et al., 2005), which suggests that dogs are able to detect most VOC. Changes in the concentrations of biogenic VOC can be used to mirror metabolic or (patho)physiological processes in the entire body. Progress in analytical chemistry in recent years has enabled the quantification and comparison of VOC of a cellular origin, for instance those released by bacterial strains such as *Pseudomonas aeruginosa* or *Streptococcus pneumonia*. VOC have been involved in the differentiation of some infectious intestinal diseases such as *Clostridium difficile, Campylobacter, Salmonella* and *Cholera*. Besa (2015) stated that VOC may be the products of different inflammatory and metabolic processes, either physiological or related to diseases of the respiratory system or other parts of the human body, or the products of oxidative stress manifested during an illness. The differences in these compounds among individuals are considerable and the concentrations may depend on several factors including metabolism, differences in lungs, and system physiology. There are many different physiological processes that affect the detectable VOC profile. Nevertheless, not all VOC released from the body are related to human metabolism. Many VOC are related to microbial or viral infections (Boots et al., 2015). They are also used to distinguish other non-infectious conditions such as the irritable bowel syndrome or the inflammatory bowel disease. Besides, volatile compounds in urine have been used to detect infections of the urinary tract and the bladder, and prostate cancer (Amann et al., 2014). The biochemical mechanisms that are behind the release of VOC related to diseases are unknown to a significant extent. No published studies have defined which VOC are detectable for dogs, and the identity of substances that dogs react to is speculative. Some studies have shown that infection-related VOC can be detected, but chemical analyses do not define the odour profile which would determine the detection behaviour of the dog (Angle et al., 2016). Several studies have shown that VOC, which we can classify as aldehydes, alcohols, alkanes, esters, fatty acids or ketones based on functional groups, may be unique for a specific patogen or infection. For instance, in a cell culture model, Schivo (2015) showed differences in the amount and representation of VOC in human tracheobronchial cells infected with the human rhinovirus. Aksenov (2012) found that VOC produced by Blymphoblastoid cells following infection with three subtypes of the influenza virus were unique for each subtype. Mashir (2018) offered an interesting finding after he had administered a live attenuated H1N1 vaccine to volunteers and proved that their exhaled breath contained a significantly higher amount of VOC in the following seven days. These studies show that there are unique VOC profiles related to virus patogens and that they can be detected in patients. This would mean they may generate a specific and characteristic odorant. Therefore, what is important for olfactory detection by dogs is the glycoproteins owing to which the surface particle may bind to specific cell receptors. There are grounds for an assumption that the replication and cell actions of the SARSCoV-2 virus may produce specific metabolites and catabolites that might be secreted by apocrine glands and generate VOC that can be detected by dogs.

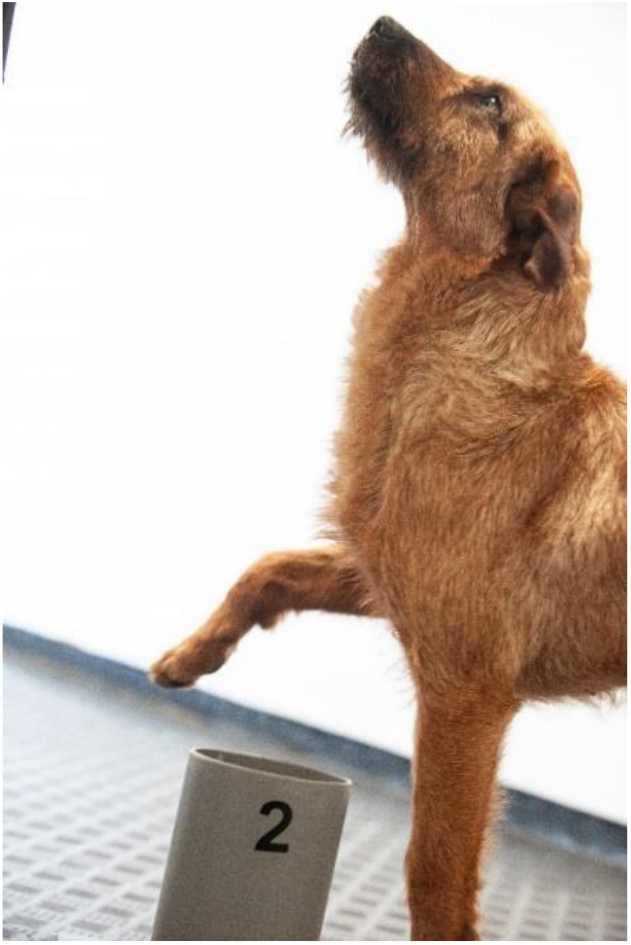

## 2 Material and methods

The pilot study involved four dogs of different ages and races. They were uncastrated males – a German Shepherd, a Giant Schnauzer, a Border Collie and a Jagdterrier. All the animals underwent previous training in different specialisations. The dogs that were trained to detect human odour (rescue training) visibly displayed behaviour adopted during this training. The German Shepherd, which underwent general training, also tended to offer behaviour adopted in his previous workplace. Therefore, animals unburdened by previous training seem to be most suitable for this specialisation. They should be properly socialised and ready to detect odours, and aged two years or older in order to ensure character stability. Castrated animals were not used in this study. It is known that primarily castrated dogs are used for similar work in the world.

The phase of imprinting Covid-positive smell (Figure 1) took place within 14 days when we already observed differences between races that were also due to previous training. Hence, only two dogs were involved in the testing phase – the Giant Schnauzer and the Jagdterrier. The health condition of the dogs was monitored throughout the training. They underwent a hematological and biochemical blood analysis. No inflammatory markers were detected in the monitored dogs. Besides, quantitative molecular detection for the presence of the SARS-CoV-2 virus was carried out in a specialised microbiological laboratory with a negative result.

**Figure 1.**
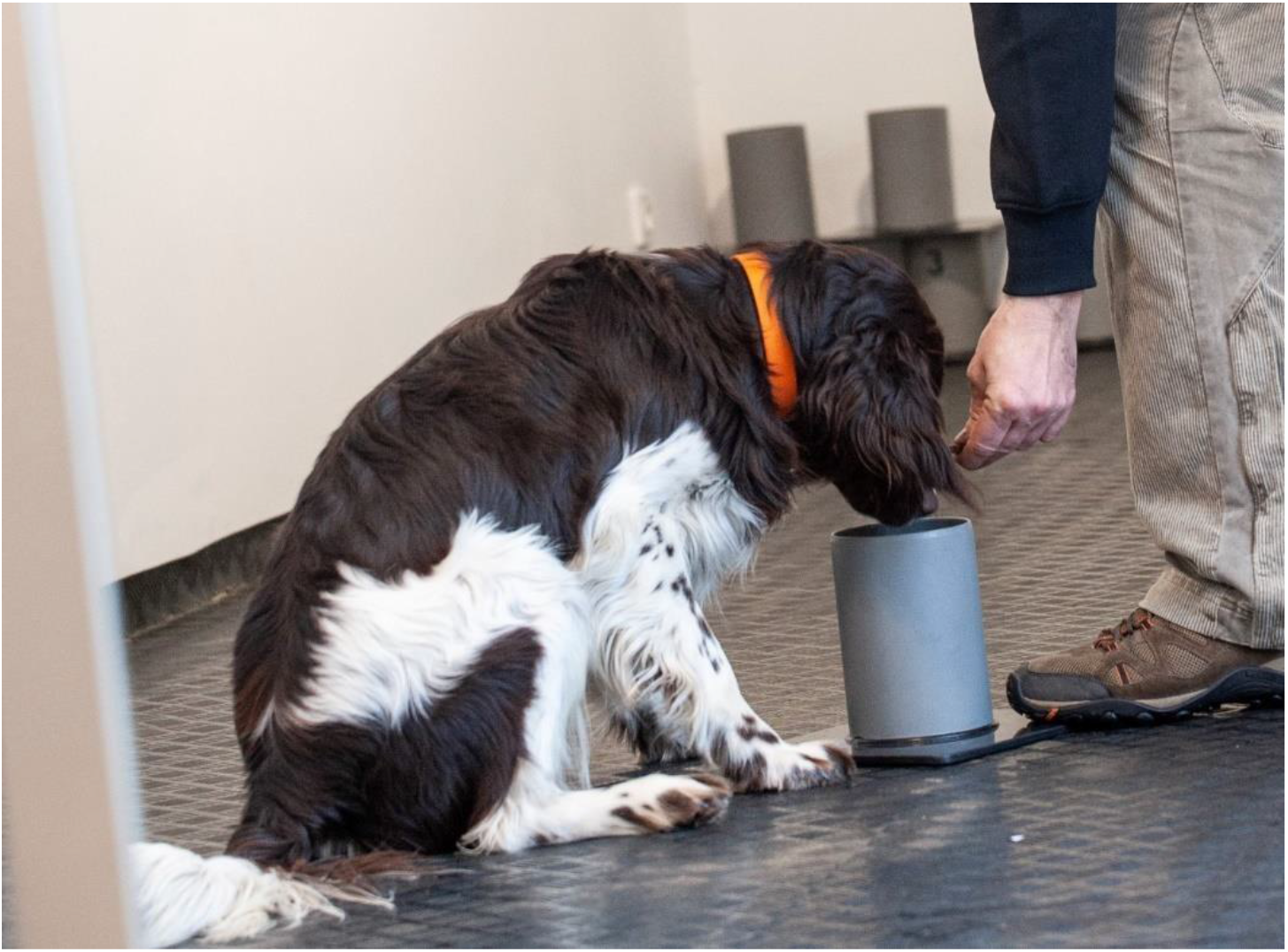
Smell imprinting phase

### Samples

For our study, we decided to use samples obtained from the body odour (torso) and breath. The sample from the upper body was chosen deliberately in order to minimise the contamination of samples imminent in other parts of the human body. The samples were placed in sampling sets. Each set comprised a zip bag, sterile surgical compresses, a sterile glass and a twist lid (Figure 2). With each person, the samples were taken in two ways: by applying the sorbent (sterile surgical compresses) onto the torso of the person and by taking a breath sample using the sorbent. The sorption time with the torso sample was set at 20 minutes. The breath sorption time was fixed at 3 minutes. The time frame for the sorption was set on the basis of findings from police practice and similar published studies. After the sorption time, the person placed the sorbent into a sterile glass and secured it with a twist lid. Because of potential contamination by other odours, the sampling set was subsequently handled exclusively in one-off latex gloves.

**Figure 2.**
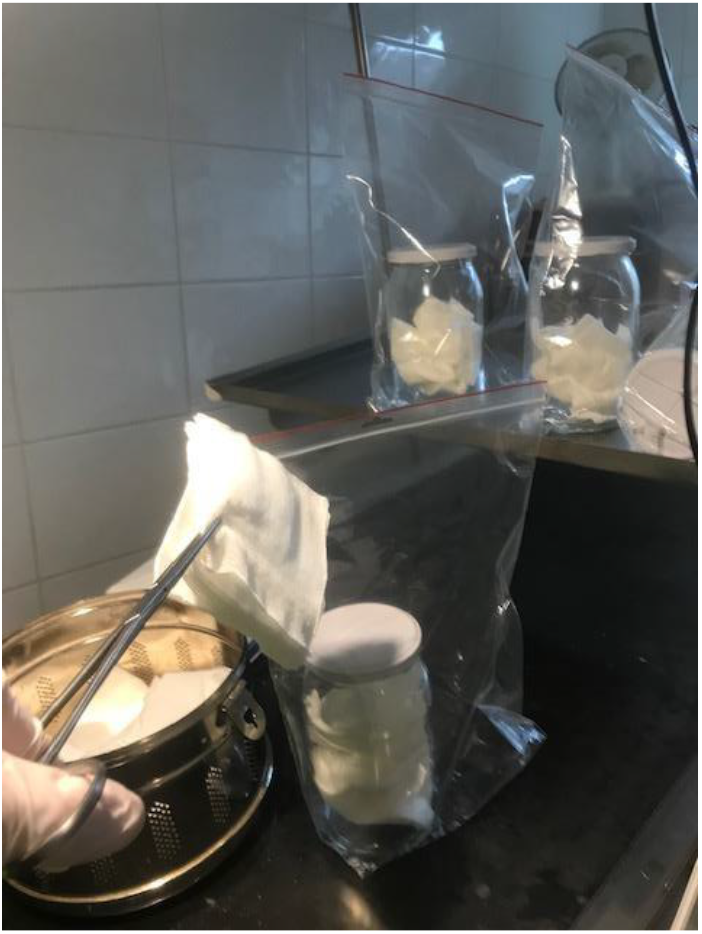
Sampling sets and manipulation

The personnel in charge of taking and handling the samples was properly trained and supplied with all necessary equipment. The donors of the odour samples were also trained, and they also filled in an anonymous study questionnaire (Appendix 1).

Positive samples were obtained from a hospital. The donors of the positive samples were diagnosed as SARS-CoV-2 positive with the help of samples obtained by means of nasopharyngeal swabs using the RT-PCR method. We collected primarily samples from donors with asymptomatic COVID-19. The donors of negative samples had no clinical symptoms related to the COVID-19 disease and they had a negative RT-PCR test for the presence of SARS-CoV-2. The set of samples included all age groups of men, women and children. The selected samples were as diverse as possible so that the only shared part of the odour signature was a change in the individual odour of the person caused by the disease. At the same time, both donor groups filled in a study questionnaire containing information for research (Appendix 1). For security’s sake, some of the positive samples were tested for the presence of the SARS-CoV-2 coronavirus at the Institute of Microbiology of the Czech Academy of Sciences. An analysis carried out by the Institute showed that only a few hours following the sorption of the odorant from the body of a Covid-positive patient, the cotton compresses no longer contained or released the RNA of the SARS-CoV-2 virus. Some published studies suggest that cotton (surgical compresses) ensures the decomposition of viral RNA within several minutes (Amann et al., 2014). The samples were used for training at least 24 hours after sorption. All samples were coded and stored in room temperature, which proved crucial for the detection abilities of the dogs. The samples were always prepared for detection in the same secure way, with the help of protective gear and sterile medical tools, in order to prevent contamination (Figure 3).

**Figure 3.**
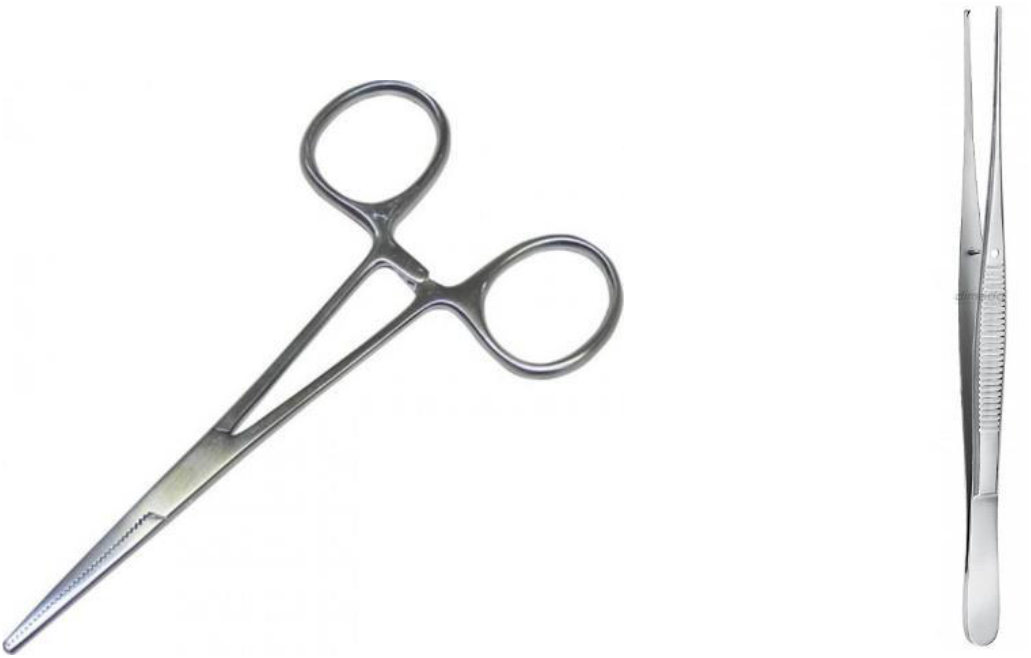
Sterile tools

The samples were crossed, that is, a sample set was blended into a target sample comprising equal shares of odorants reflecting the age and sex. Each crossed sample, which was used in both the training and testing phases, comprised samples from group of different donors. In the training phase, the dogs had the samples at their disposal three times in a row at the most. In the testing phase, they were always expected to detect a new Covid-positive target sample in each battery. In this study, we have used a total of 228 samples, of which 156 positive and 72 negative for the presence of the SARS-CoV-2 coronavirus.

### Experiment and results

Two specially trained detection dogs were involved in the first experiment. Dog 1 was a Giant Schnauzer, an uncastrated six-year-old male. Dog 2 was a Jagdterrier, a seven-year-old uncastrated male. Both dogs successfully passed a two-week phase of imprinting the target odorant. Subsequently, they were capable of comparing the samples. After the training phase, we decided to test the two detection dogs. The tests followed identical protocolar techniques. The dogs faced a row of six adjusted detection tubes (Figure 4) containing the target sample of the odorant of a Covid-positive donor, a negative sample from a Covid-negative donor, and so-called fake samples, which are neutral and designed to distract the detection dogs.

**Figure 4.**
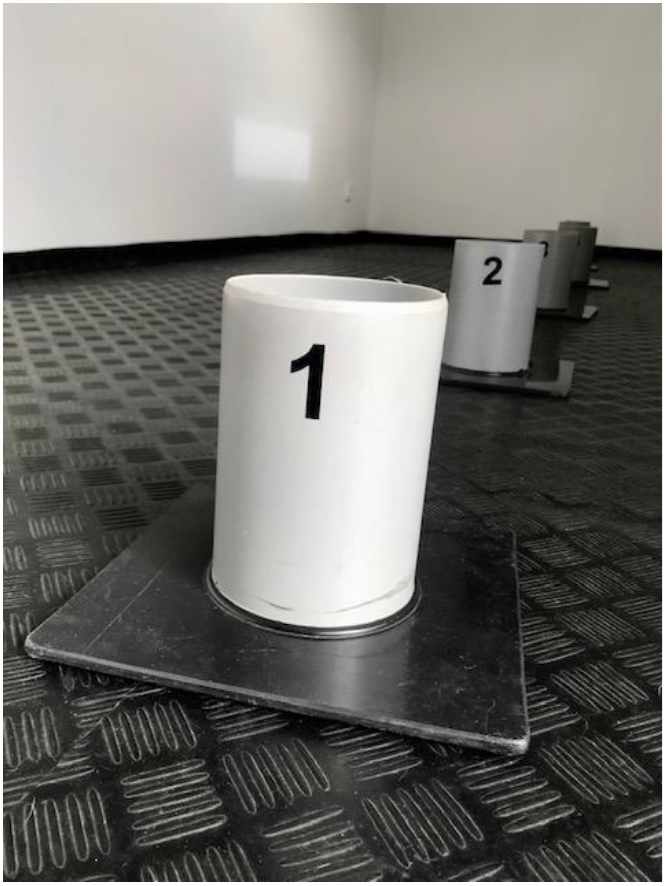
Detection tubes

**Figure 5.**
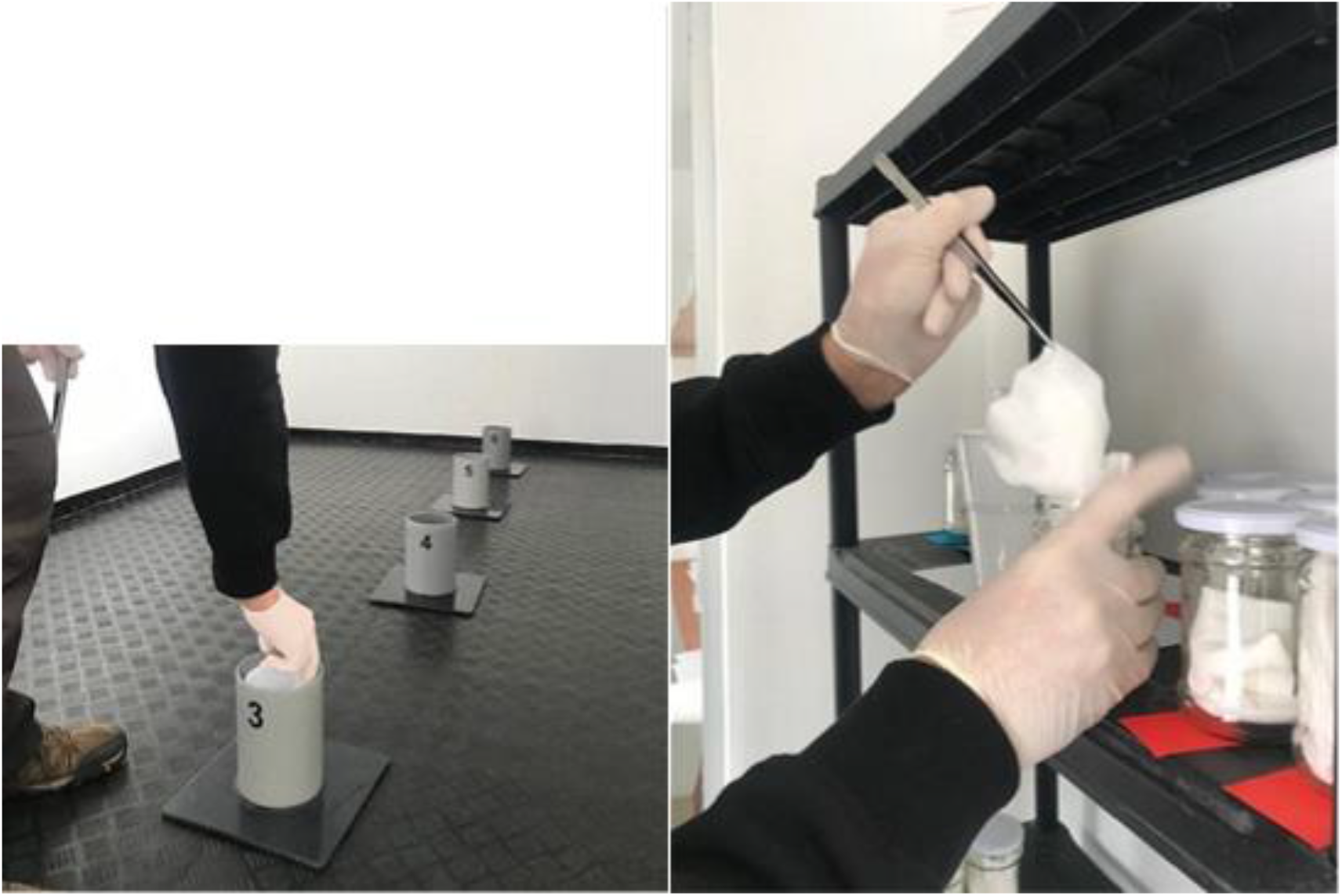
Preparing the samples

**Figure 6.**
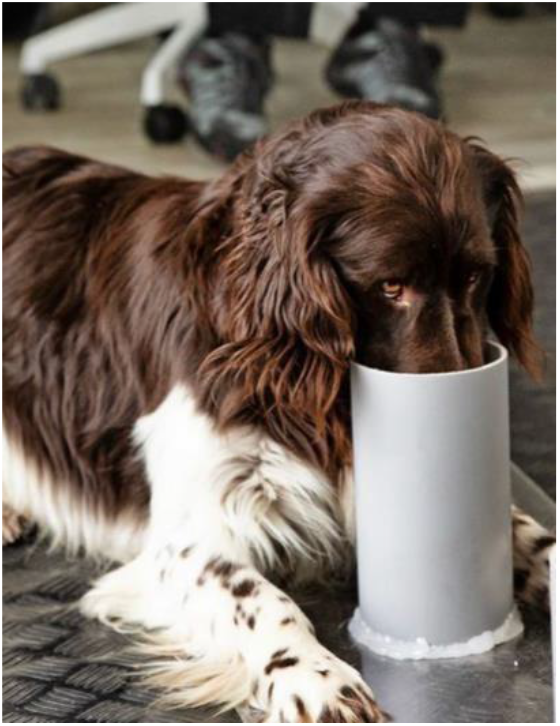
Identifying a positive sample

Each testing battery comprised the same representation of samples, that is, one Covid-positive sample, one Covid-negative sample and four neutral samples. It was only the order of the samples that changed for each tested dog. The coach never knew the composition and placement of the samples. He was warned by an acoustic signal when the detection was correct. The dogs received a standard reward for detecting the sample correctly. Out of a total number of 100 detected samples, the dogs correctly identified the positive samples in 95 cases. Of the total number of 100 detected negative samples, they only identified six. The reasons why the dogs did not identify the positive samples but the negative ones are subject to further research. Unfortunately, it was not possible to subject the donors of samples that were detected erroneously to another PCR test. All experiments were carefully registered and recorded to enable a further analysis.

**Table 1:** Detection dog results

| Dog name | Detected by dog | SARS-CoV-2 infection status |  | Total | Diagnostic specificity (Sp) | Diagnostic sensitivity (Se) | Negative predictive value (NPV) | Positive predictive value (PPV) |
| --- | --- | --- | --- | --- | --- | --- | --- | --- |
|  |  | Negative | positive |  |  |  |  |  |
| Renda | No | 48 | 3 | 100 | 0,96 | 0,94 | 0,94117647 | 0,95918367 |
|  | Yes | 2 | 47 |  |  |  |  |  |
| Laky | No | 46 | 2 | 100 | 0,92 | 0,96 | 0,95833333 | 0,92307692 |
|  | Yes | 4 | 48 |  |  |  |  |  |
| Total | No | 94 | 5 | 100 | 0,94 | 0,95 | 0,94949495 | 0,94059406 |
|  | Yes | 6 | 95 |  |  |  |  |  |

**Table 2:**
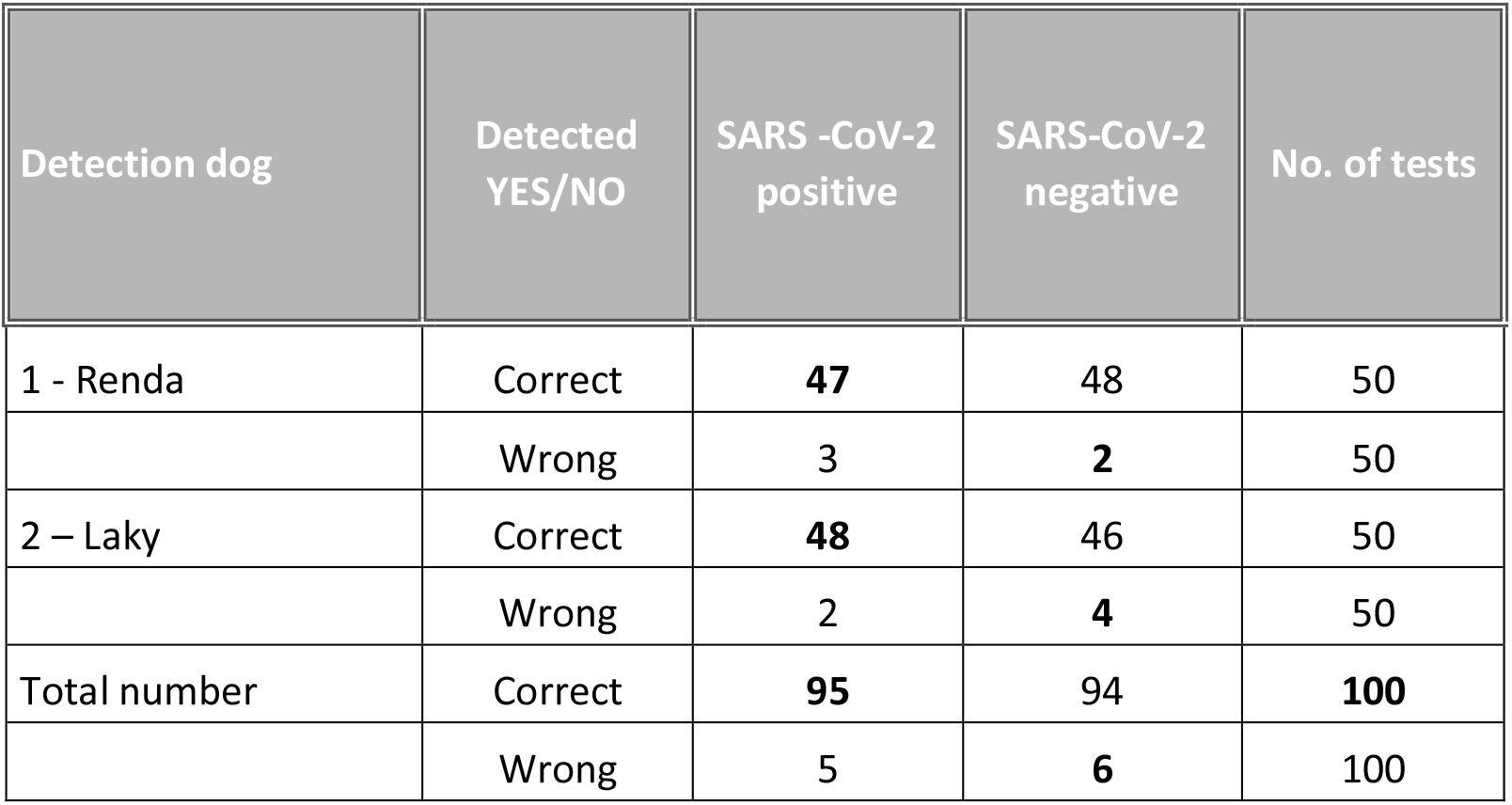
Detection dog results

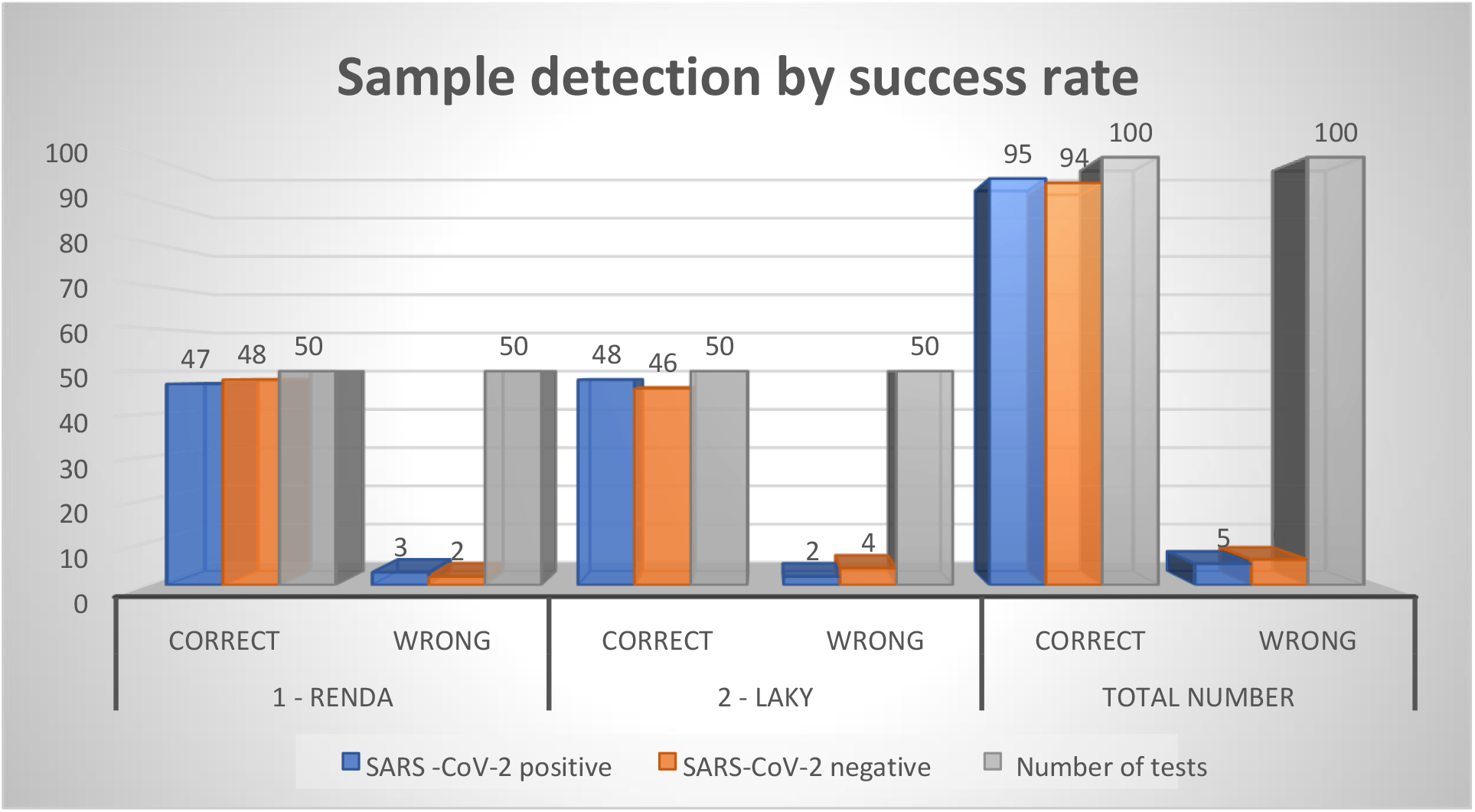

CORRECT WRONG TOTAL NUMBER

SARS-CoV-2 positive SARS-CoV-2 negative number of tests

## 3 Discussion

A timely and precise detection of individuals infected with the SARS-CoV-2 coronavirus is one of the key factors in battling the pandemic. Data from our pilot study suggest that specially trained dogs could become a convenient detection tool for population screening owing to a relatively high sensitivity of 95% and specificity of 94%. For instance, fast diagnostic tests detecting antigens (Lepu) reach a sensitivity of 45.5% and specificity of 89.2% (Baro et al., 2021). Therefore, these antigen tests are obviously not suitable for virus detection in asymptomatic individuals. Cohen (2020) says that the SARS-CoV-2 coronavirus diagnostics using the RT-PCR method by means of a nasopharyngeal swab may reach a false detection rate of up to 25%.

Our study worked with samples from patients who were asymptomatic. The control negative samples were from healthy individuals who had no symptoms of a respiratory disease and who had negativity confirmed by an RT-PCR method. With the false positive detection by dogs, we cannot determine with any exactitude whether this may have been for instance a case of cross detection with dogs reacting to another human coronavirus.

To resolve these problems, we will need subsequent studies targeting the differentiation of other respiratory infections, stages of the disease, the detection of pre-symptomatic patients, etc. Nevertheless, the results of our pilot study and of foreign studies suggest there are unique VOC imprints in the odour signature of a COVID-19-positive individual, which can be detected by dogs.

## 4 Conclusion

The pilot study suggests that trained detection dogs are able to distinguish and subsequently identify samples of an odorant from individuals infected with the SARS-CoV-2 coronavirus with a high degree of sensitivity (95%) and specificity (94%). Dogs trained in this way could be a reliable biological detector of the COVID-19 disease. The detection method seems to offer a suitable alternative for instance in countries with restricted access to commonly used diagnostic tests or to the testing of child and senior populations. From the operational point of few, detection dogs can be a top-quality biological detector offering a highly sensitive sensoric on-site system in real time, without the need to collect, process or analyse samples, which provides certain advantages against machines. Recommendations stemming from this pilot study are aimed at subsequent research, especially when it comes to the differentiation of other virus diseases or the use of more suitable methods of odorant sorption. Further work is needed to better understand the detection abilities of dogs.

## Data Availability

All data referred to in the manuscript we have in secure storage without direct access from Internet

## Acknowledgements

MUDr. Jan Třešňák; Mudr. Markéta Švábová; MUDr. Soňa Peková, PhD.; MUDr. Jan Laryš; MVDr. Blanka Tešnarová; RNDr. Kateřina Bíšová; Czech Red Cross; Thomayer University Hospital; Olomouc University Hospital; Getxent and the Prague Fire Brigade

